# The role of Mediterranean diet adherence, smoking and their interactions in epigenetic age acceleration: A cross-sectional analysis of the Airwave cohort

**DOI:** 10.64898/2026.06.21.26355777

**Authors:** Azaan Zaki, Chung-Ho E. Lau, Rebeca Eriksen, Gary Frost, Ian Mudway, Oliver Robinson

**Author notes:** **Correspondence to**: Azaan Zaki; **email**. Joint senior authors (Azaan Zaki, Oliver Robinson).

## Abstract

**Background:** Epigenetic clocks are markers of biological aging that may vary in their sensitivity to environmental stressors and lifestyle modifiers. To evaluate the utility of these biomarkers as sensors of the human exposome, we investigated how they respond to two powerful and opposing exposures: smoking, a source of oxidative stress, and the antioxidant-rich Mediterranean diet.

**Objectives:** We assessed the sensitivity of eleven epigenetic clocks to diet and smoking and evaluated whether Mediterranean diet adherence modifies associations between smoking and epigenetic aging.

**Methods:** We analysed 928 participants (mean age 41 years, 59% male) from the Airwave Health Monitoring Study. Linear regression models assessed associations between Mediterranean Diet Score (MDS) and epigenetic age acceleration (EAA), alongside smoking status and blood cotinine. Interaction terms between smoking status and MDS were included to detect dietary attenuation of smoking-related EAA. Models were adjusted for demographic, socioeconomic, lifestyle, and psychological covariates.

**Results:** Higher MDS was associated with lower EAA for GrimAge (β = −0.07 SD; 95% CI: −0.13, −0.01) and Bernabeu (β = −0.08 SD; 95% CI: −0.14, −0.02) after false discovery rate correction. Smoking was strongly associated with increased EAA, particularly for GrimAge, Bernabeu, and DunedinPACE. Among current smokers, effect sizes were greater in those with lower dietary adherence (e.g. GrimAge: 1.79 SD, 95% CI: 1.54, 2.04) compared with those with higher adherence (1.35 SD, 95% CI: 1.01, 1.68; P__interaction_ < 0.001). Similar attenuation patterns were observed for Bernabeu. Higher intake of fruits, vegetables, and whole grains contributed most to the attenuation of smoking-related EAA.

**Conclusions:** Our findings indicate that certain epigenetic clocks effectively capture the tension between harmful and protective exposures within the exposome. Rather than suggesting that diet neutralises the risks of tobacco, these results demonstrate that specific clocks are sensitive enough to monitor how lifestyle factors modify molecular responses to environmental toxins. This highlights the value of second-generation clocks in quantifying biological resilience.

## INTRODUCTION

Aging is the most important risk factor for most chronic diseases, physical and cognitive impairment, and death [1]. It has therefore been proposed that targeting aging as a biological process, may represent a strategy to forestall age-associated disease [2]. As life expectancy continues to grow, the promise of therapeutic targeting of aging to extend healthy lifespan has come into sharper focus. To achieve this goal, there is growing interest in quantifying biological age, as a biomarker for chronic disease risk [3].

Epigenetic clocks have been developed using different training strategies, which may influence their sensitivity to environmental and lifestyle exposures. First-generation clocks were trained to predict chronological age[4, 5], whereas later-generation clocks incorporate biomarkers of physiological function, mortality risk, or longitudinal aging processes. Among these, mortality-trained clocks include PhenoAge, derived by constructing a composite “phenotypic age” from clinical biomarkers and training a methylation predictor of that phenotype [6], and GrimAge and the Bernabeu clock, which were developed using DNA methylation surrogates of circulating proteins and smoking pack-years to predict mortality risk [7, 8]. Other approaches aim to capture specific biological processes. DNAmTL and EpiTOC estimate telomere length and mitotic history, respectively, and may be considered ‘mechanistic’ clocks [9, 10]. In contrast, pace-of-aging clocks, such as DunedinPACE, quantify the rate of physiological decline using longitudinal biomarker data, providing a dynamic measure of aging [11]. More recently, causality-informed clocks have been developed using genetic causal inference; for example, CausAge was constructed using CpG sites identified through epigenome-wide Mendelian randomization, with complementary measures capturing detrimental (DamAge) and adaptive (AdaptAge) aging processes [12].

Compared to first-generation clocks, these later-generation approaches integrate a broader range of biological pathways, including metabolic, inflammatory, and lipid-related processes, which are influenced by lifestyle and environmental exposures such as diet, stress, and physical activity [13]. An overview of these clocks is provided in **Table S1**. Whilst the use and variety of these biological clocks has expanded rapidly, there remains a need to evaluate and compare their performance if they are to be widely deployed for health surveillance. Some attempts have been made to do this [14], but overall the results of these studies have been equivocal [3], with few studies using standardized and equivalent measurement units to compare clocks across single, or multiple cohorts [15, 16]. There is currently no consensus on the methods for evaluating the sensitivity of these clocks beyond the primary criterion that they should be age sensitive.

Diet is a key modifiable determinant of healthy ageing, yet its relationship with epigenetic ageing biomarkers remains incompletely characterised. Among dietary patterns, the Mediterranean diet (MD) is of particular interest because adherence to this pattern has been associated with improved survival and healthier ageing trajectories [17, 18]. More recent work has also shown that higher overall diet quality is associated with decelerated epigenetic ageing [19]. Mechanistically, dietary exposures may influence ageing-related pathways through effects on metabolic health, oxidative stress, inflammation, and nutrient sensing. For example, flavonoids and other antioxidant-rich dietary components may improve glycaemic regulation and reduce oxidative damage [20], while fibre-rich whole grains support metabolic health [21]. These mechanisms are relevant to biological ageing because disrupted insulin signalling, adiposity, and metabolic dysfunction are closely linked to ageing-related processes [22], and higher-quality carbohydrate intake has also been associated with healthy ageing outcomes [23]. Whilst indices such as the Dietary Inflammatory Index are designed to quantify the inflammatory potential of diet based on associations with specific immune and inflammatory biomarkers, the Mediterranean diet offers a validated and holistic representation of a lifestyle pattern characterised by high antioxidant capacity. It therefore serves as a robust model to investigate whether epigenetic clocks can detect the cumulative protective effects of a complex nutritional exposure against environmental stress.

In contrast, cigarette smoking is among the most potent and well-characterized lifestyle risk factors for premature morbidity and mortality, reducing life expectancy by a decade [23]. Smoking accelerates ageing-related processes across multiple physiological systems, with clear links to cardiopulmonary morbidity [24–26], chronic obstructive pulmonary disease (COPD) and lung cancer [27–31]. In addition there is sufficient evidence smoking increases oxidative stress and inflammation [32], as well as telomere attrition [33]. Because these processes overlap closely with the biological domains captured by epigenetic clocks, smoking provides a useful environmental ageing stressor against which the sensitivity of these biomarkers can be evaluated. Smoking cessation is known to reduce cardiovascular mortality, diminish lung cancer risk, and improve health outcomes at any age [34–36], and while smoking-associated DNA methylation changes may be partially reversible, evidence for reversal of epigenetic ageing itself remains limited. Within this framework, smoking serves as a high-contrast environmental stressor. It provides a rigorous context in which to evaluate whether epigenetic clocks can capture the biological tension between harmful exposures and protective lifestyle factors.

We examined the associations of smoking and Mediterranean diet adherence with epigenetic age acceleration across eleven clocks in the Airwave Health Monitoring Study, an occupational cohort of British police employees [37]. We used multiple indices of exposure, including self-reported status, time since quitting and blood cotinine, to strengthen inference beyond subjective reporting. MD adherence was assess using a validated 7-day food diary This working age cohort represents a relevant life stage where lifestyle-related changes in biological ageing may be detected prior to the onset of overt disease. We hypothesised that smoking-related age acceleration would be most evident in later-generation clocks and that greater Mediterranean diet adherence would be associated with lower age acceleration and a measurable attenuation of the smoking-related ageing signal.

## METHODS

### Study population

This analysis used data from the Airwave Health Monitoring Study, an occupational cohort of ∼51,000 police employees in Great Britain. Full recruitment and data collection procedures are described elsewhere [37]. DNA methylation profiling was conducted on a subset of 1,129 participants, selected based on the availability of high-quality DNA, genotyping, and metabolomics data. Quality control exclusions were applied for bisulfite conversion failure, low probe call rate, and gender mismatch, yielding 1,115 participants with high-quality methylation data. These were subsequently merged with dietary questionnaire data collected between 2007 and 2012. Only 928 participants had complete data for both methylation and diet variables and were included in the final analysis. Details of the study design and analyses is referred to in **Fig. S1**.

All participants provided written informed consent, and the study received ethical approval from the NHS Multi-Site Research Ethics Committee (MREC/13/NW/0588).

### Dietary assessment

Dietary intake was evaluated using a 7-day estimated-weight food diary. Participants were provided with instructions, including portion size guidance based on food portion photographs and common household measurements, alongside a general questionnaire to validate the recorded intake. This diary method had been previously validated in a larger UK epidemiological study [38]. Participants submitted their completed diaries either by post or during their health screening visit at the clinic. The data were analysed using Dietplan (version 6.0; Forestfield Software), which incorporates the UK nutrient database based on *McCance and Widdowson’s The Composition of Foods* (2008, 6th edition, published by the UK Food Standards Agency). A study-specific operational manual and quality control protocol were developed to standardize the coding process and ensure the accuracy of the food diary data [39]. To evaluate potential bias due to misreporting of dietary intake, energy reporting plausibility was assessed using the Goldberg method. Reported energy intake was compared with estimated basal metabolic rate (BMR), calculated using Schofield equations based on age, sex, and body weight [40]. Cut-offs for plausible reporting incorporated within-person variation in energy intake, error in BMR estimation, and variation in physical activity level, assuming a 7-day dietary assessment period. Participants were classified as under-reporters or plausible reporters based on established criteria.

The MDS was calculated to assess adherence to the Mediterranean dietary pattern using key dietary components. Participants’ intakes of healthy food groups, including fruits, vegetables, legumes, fish, nuts, and whole grains, were scored as 1 if they exceeded the sex-specific median intake and 0 otherwise. Intakes of unhealthy food groups, such as red meat and total dairy, were scored as 1 if they were below the sex-specific median and 0 if they exceeded it. Moderate alcohol consumption was scored as 1 for men consuming 10–50 grams/day and women consuming 5–25 grams/day, with scores of 0 assigned for intakes outside these ranges. The ratio of monounsaturated to saturated fatty acids (MUFA: SFA), a proxy for olive oil consumption, was also included, with a score of 1 assigned if it exceeded the sex-specific median and 0 otherwise. The total MDS, ranging from 0 to 10, was obtained by summing all component scores, where higher values indicate greater adherence to the Mediterranean diet [17, 18].

### Smoking exposure

Smoking measures included smoking status (n = 928), blood cotinine (n = 823), and time since quitting in years among former smokers (n = 227). Smoking status was categorised as current, former, or never smoker, based on self-reported questionnaires. Cotinine levels, an objective biomarker of recent nicotine exposure [41], were measured in blood samples using liquid chromatography–mass spectrometry (LC-MS), a highly sensitive and specific method for detecting nicotine metabolites, as previously described [42, 43]. The assay provided relative cotinine values, which were normalized to account for batch effects or variations in sample processing. Cotinine values were used to cross-validate self-reported smoking status (**Fig. S3**). Concordance between biochemical and self-reported smoking categories was high across groups, supporting the validity of self-report data. Blood cotinine was measured for all 928 participants. Of these, 823 had non-zero cotinine values, while 105 self-reported never smokers had cotinine values of zero, consistent with their reported smoking status. Analysis of cotinine and epigenetic clocks was conducted only in participants with detectable cotinine levels.

### Covariates

Socio-demographic, health and lifestyle data were collected via a self-administrated electronic questionnaire, which the participants filled in during their clinic visit. Variables used for the present study included: age, sex, BMI, smoking status, diet quality, household income, education level, alcohol consumption, anxiety levels, and physical activity. Models examining Mediterranean Diet Score, individual Mediterranean diet components, or diet–smoking interaction terms were additionally adjusted for total energy intake. Covariates were defined as follows: Household income was categorized into three levels: low (<£38,000), medium (£38,000-£77,999), and high (>£78,000). Education level was classified into low (GCSE/O-Level/CSE or equivalent, vocational qualifications), medium (A-Levels/Highers or equivalent), and high (bachelor’s degree or postgraduate qualifications). Alcohol consumption was divided into three categories: non-drinker (no alcohol use), occasional drinker (≤14 alcohol units/week for women and ≤21 units/week for men), and habitual drinker (>14 units/week for women and >21 units/week for men) [44]. BMI was classified as normal weight (<25 kg/m²), overweight (25 ≤ BMI < 30 kg/m²), or obese (BMI ≥ 30 kg/m²) [45].

Physical activity was measured using the International Physical Activity Questionnaire short version and the metabolic equivalent minutes per week were calculated for each participant and categorised; high (at least 60 min/d of at least moderate-intensity activity), moderate (at least 30 min/d of at least moderate-intensity activity), low (no activity is reported or less then medium category) [46]. Anxiety was assessed using the anxiety subscale of the Hospital Anxiety and Depression Scale (HADS-A), with scores categorized as normal (0-7), borderline abnormal (8-10), and abnormal (11-21) [47].

### Epigenetic age assessment

DNA methylation data acquisition and preprocessing have been described in detail elsewhere [48]. Briefly, DNA was extracted from peripheral blood cells, and samples with insufficient DNA concentration (<25 ng/µl) were excluded. For methylation profiling, 500 ng of DNA per sample underwent bisulfite conversion (EZ DNA Methylation-Lightning™ Kit, Zymo Research, Orange, CA) and was hybridized to the Infinium HumanMethylationEPIC BeadChip. Preprocessing included quality control using SNP genotyping, background subtraction, dye-bias correction, and outlier removal based on BeadChip control intensities. DNA methylation levels were quantified as β-values and used in all statistical analyses.

DNAm-based epigenetic ages (Horvath, Hannum, PhenoAge, GrimAge, DNAmTL[4–7, 9]) were calculated using the online DNAmAge calculator [49] from normalised DNAm data and crosschecked via the Bioconductor package Methylclock (version 0.5.0). The GrimAge algorithm incorporates DNAmPACKYRS, which is largely driven by methylation at smoking-associated loci such as cg05575921 in the AHRR gene. Bernabeu age [8], AdaptAge, DamAge, and CausAge [12], EpiTOC [10] and DunedinPACE [11] were calculated using published coefficients outlined from the original papers.

Age acceleration estimates were obtained from these raw epigenetic clock estimates by extracting the residuals from the regression model that regresses predicted age on chronological age – HorvathAA, HannumAA, PhenoAA, GrimAA, DNAmTLAdjAge, BernabeuAA, AdaptAA, DamAA, CausAA. All clock estimates (age acceleration, DunedinPACE & EpiTOC) were standardized by rescaling them to have a mean of 0 and a standard deviation of 1, to enable comparability across clocks. DNAmTLAdjAge was multiplied by -1 to align effect directionality with other clocks, such that higher values represent greater age acceleration.

### Statistical Analysis

Continuous predictors, including the MDS, blood cotinine levels, and time since quitting (in years), were also standardised (mean = 0, SD = 1) to allow interpretation of associations per SD increase in the exposure variable.

Associations between MDS and smoking measures and each epigenetic clock were assessed using linear regression models for each exposure with EAA (or corresponding clock output) as the outcome variable, adjusting for key covariates selected based on a constructed Directed Acyclic Graph (DAG) (**Fig. S2**) derived from the literature. These include age, sex, BMI, household income, education level, alcohol consumption, physical activity, and anxiety levels. MDS was additionally included as a covariate for analyses with smoking exposures, while smoking status was included as a covariate for analyses with MDS. Total energy intake was additionally included in models where Mediterranean diet score, individual dietary components, or diet–smoking interaction terms were examined.

To evaluate whether diet quality modified the association between smoking and EAA, participants within each smoking status group were divided into high and low Mediterranean diet adherence groups based on the cohort-wide median value, and differences in EAA were examined within each stratum. Next, we included an interaction term between smoking status (categorical: never, former, current) and MDS (continuous) in fully adjusted linear regression models to formally test whether the association between smoking and EAA varied by diet adherence. Interaction analyses were prioritised for epigenetic clocks demonstrating the consistent associations across smoking exposures, as effect modification is most interpretable in the presence of a clear exposure–outcome relationship. Finally, to explore which dietary components were driving any observed interaction effects, we conducted additional interaction analyses between smoking status and individual components of the MDS (e.g., fruit, vegetables, fish, whole grains, dairy), tested separately for each component, and evaluated their moderating effect on smoking-related EAA.

In sensitivity analysis, associations between Goldberg-defined reporting status (under-reporter vs. plausible reporter) and selected epigenetic ageing measures (GrimAge, BernabeuAA and DunedinPACE) were evaluated using linear regression models adjusted for age, sex and BMI.

Multiple testing correction was conducted using Benjamini–Hochberg False Discovery Rate FDR [50]. All analyses were performed in R version 4.3.1.

## RESULTS

### Study Population

**Table 1** shows the descriptive characteristics of the subset of 928 study participants with available DNA methylation data from the Airwave cohort. The cohort’s average age is 41.0 years (SD = 9.3), and BMI is 27.2 kg/m² (SD = 4.4), with 96.9% being White and 41.2% female. These characteristics closely align with the full AIRWAVE Health and Monitoring Study cohort [37]. Education level, alcohol consumption, and anxiety significantly differed across smoking groups (p < 0.001, p = 0.006, p = 0.003, respectively). A higher percentage of never smokers (30.5%) had a high education level compared to smokers (15.3%) and former smokers (22.9%). Current smokers had the highest percentage of habitual drinkers (49.0%), and the highest proportion of those with abnormal anxiety levels (14.3%). Relative cotinine levels were significantly higher in current smokers compared to former and never smokers (p < 0.001) (**Fig. S3**). The Mediterranean Diet score (MDS) was significantly lower in current smokers (mean = 4.3) compared to former smokers (mean = 4.7) and never smokers (mean = 4.9), with p = 0.002 (**Fig. S4**). Based on the Goldberg method [51], 44.7% of participants were classified as under-reporters and 55.3% as plausible reporters, with no substantial over-reporting observed.

**Table 1.** Baseline descriptive characteristics of AIRWAVE cohort stratified by smoking status (N = 928).

| Characteristic |  | Current Smoker<br>(N = 98) | Former Smoker<br>(N = 227) | Never Smoker<br>(N = 603) | Total<br>(N = 928) | p-value |
| --- | --- | --- | --- | --- | --- | --- |
| **Age (Y)** | Mean (SD) | 42.0 (10.0) | 43.4 (9.2) | 40.0 (9.0) | 41.0 (9.3) | <0.001 |
| **Gender** | Female % | 38 (38.8) | 103 (45.4) | 242 (40.1) | 383 (41.2) | 0.431 |
| **Ethnicity** | White % | 91 (92.9) | 216 (95.2) | 592 (98.2) | 899 (96.9) | 0.004 |
| **Relative blood cotinine (arbitrary units)** | Mean (SD) | 1.47 (1.12) | 0.005 (0.03) | 0.001 (0.00) | 0.16 (0.58) | <0.001 |
| **Household Income** | High % | 9 (9.2) | 20 (8.8) | 66 (10.9) | 95 (10.2) | 0.794 |
|  | Medium % | 57 (58.2) | 143 (63.0) | 369 (61.2) | 569 (61.3) |  |
|  | Low % | 32 (32.7) | 64 (28.2) | 168 (27.9) | 264 (28.4) |  |
| **Education level** | High | 15 (15.3) | 52 (22.9) | 184 (30.5) | 251 (27.1) | <0.001 |
|  | Medium | 29 (29.6) | 66 (29.1) | 191 (31.7) | 286 (30.8) |  |
|  | Low | 54 (55.1) | 109 (48.0) | 228 (37.8) | 391 (42.1) |  |
| **Alcohol Consumption** | Habitual Drinker % | 48 (49.0) | 92 (40.5) | 118 (19.6) | 258 (27.8) | 0.006 |
|  | Occasional Drinker % | 38 (38.8) | 108 (47.6) | 192 (31.8) | 338 (36.4) |  |
|  | Never Drinker % | 12 (12.2) | 27 (11.9) | 305 (50.6) | 344 (37.1) |  |
| **BMI** | Mean (SD) | 27.0 (3.7) | 27.7 (4.9) | 27.0 (17.6) | 27.2 (4.4) | 0.108 |
| **Physical Activity** | High % | 12 (12.2) | 15 (6.6) | 50 (8.3) | 77 (8.3) | 0.148 |
|  | Moderate % | 10 (8.6) | 13 (5.7) | 47 (7.8) | 70 (7.5) |  |
|  | Low % | 76 (77.6) | 199 (87.7) | 506 (83.9) | 781 (84.2) |  |
| **Anxiety** | Abnormal % | 14 (14.3) | 22 (9.7) | 40 (6.6) | 76 (8.2) | 0.003 |
|  | Borderline Abnormal % | 10 (10.2) | 45 (19.8) | 92 (15.3) | 147 (15.8) |  |
|  | Normal % | 74 (75.5) | 160 (70.5) | 471 (78.1) | 705 (76.0) |  |
| **Med Diet score** | Mean (SD) | 4.3 (1.8) | 4.7 (1.8) | 4.9 (1.8) | 4.8 (1.8) | 0.002 |
| **Q1 (Lowest)** |  | 54 (55.1) | 109 (48.0) | 254 (42.1) | 417 (44.9) |  |
| **Q2** |  | 16 (16.3) | 41 (18.1) | 129 (21.4) | 186 (20.0) |  |
| **Q3** |  | 14 (14.3) | 39 (17.2) | 97 (16.1) | 150 (16.2) |  |
| **Q4 (Highest)** |  | 14 (14.3) | 38 (16.7) | 123 (20.4) | 175 (18.9) |  |

Variable categorisation
Household income - Low: Income less than 38000, Medium: Income between 38000 and 77999, High: Income more than 78000
Education level - Low: GSCE/O-Level/CSS, left school before taking O levels / GCSEs, Vocational qualifications (NVQ1+2). Medium: A levels / Highers or equivalent (NVQ3). High Education Level: Bachelor's degree or equivalent (NVQ4), Postgraduate qualifications
\*Alcohol Consumption - non-drinker - No alcohol use, occasional drinker: ( $\leq 14$ alcohol units/week for women and $\leq 21$ alcohol units/week for men) or habitual drinker ( $> 14$ alcohol units/week for women and $>21$ alcohol units/week for men)
Physical Activity - Low: Does not meet criteria for moderate or high activity, Moderate - 5 or more days of moderate-intensity activity or walking of at least 30 minutes per day. High: 7 or more days of any combination of walking, moderate-intensity, or vigorous-intensity activities achieving a minimum of at least 3000 MET-minutes/week. Scoring protocol followed the International Physical Activity Questionnaire (https://sites.google.com/view/ipaq/score)
Anxiety - Probable anxiety was measured using the anxiety subscale of the HADS-A. Normal: 0-7 Borderline abnormal (borderline case): 8-10 Abnormal (case) 11-21.
Values are presented as mean (SD) for continuous variables and n (%) for categorical variables. P-values compare current, former, and never smokers and were calculated using one-way analysis of variance (ANOVA) for continuous variables and Pearson's chi-square tests for categorical variables.

### Epigenetic clock performance

The performance of each epigenetic clock in relation to chronological age, including correlation coefficients, is presented in the Supplement (**Fig. S5**). Horvath and Hannum had strong positive correlations with chronological age (ρ = 0.91 and ρ = 0.92, respectively; both p < 0.001). DNAmTL showed a moderate negative correlation (ρ = -0.52, p < 0.001). PhenoAge and GrimAge exhibited strong positive correlations (ρ = 0.91, p < 0.001; ρ = 0.89, p < 0.001) with chronological age in the AIRWAVE cohort. GrimAge and PhenoAge are scaled in units of years. DunedinPACE and EpiTOC had weak positive correlations (ρ = 0.26, p < 0.001; ρ = 0.34, p < 0.001). Bernabeu showed a strong positive correlation (ρ = 0.90, p < 0.001). Moderate positive correlations were found for AdaptAge (ρ = 0.31, p < 0.001), while DamAge and CausAge showed strong positive correlations (ρ = 0.72, p < 0.001; ρ = 0.85, p < 0.001).

### Mediterranean diet and EAA

The analysis revealed inverse associations between MDS and EAA for a subset of clocks (**Fig. 1A, Table S2**). Each standard deviation increase in MDS was significantly associated with lower EAA as measured by GrimAge (–0.07 SD, 95% CI: –0.13, –0.01, FDR p = 0.033) and BernabeuAA (–0.08 SD, 95% CI: –0.14, –0.02, FDR p = 0.033). Nominal inverse associations were also observed for EpiTOC (–0.07 SD, 95% CI: –0.13, 0.00), DunedinPACE (–0.07 SD, 95% CI: –0.13, –0.01), and DNAmTLAdjAge (–0.06 SD, 95% CI: – 0.13, 0.00), but these did not remain significant after multiple testing correction (all FDR-adjusted p-values > 0.05). No clear associations were observed for the Horvath, Hannum, PhenoAge, or causality-enriched clocks.

**Figure 1.**
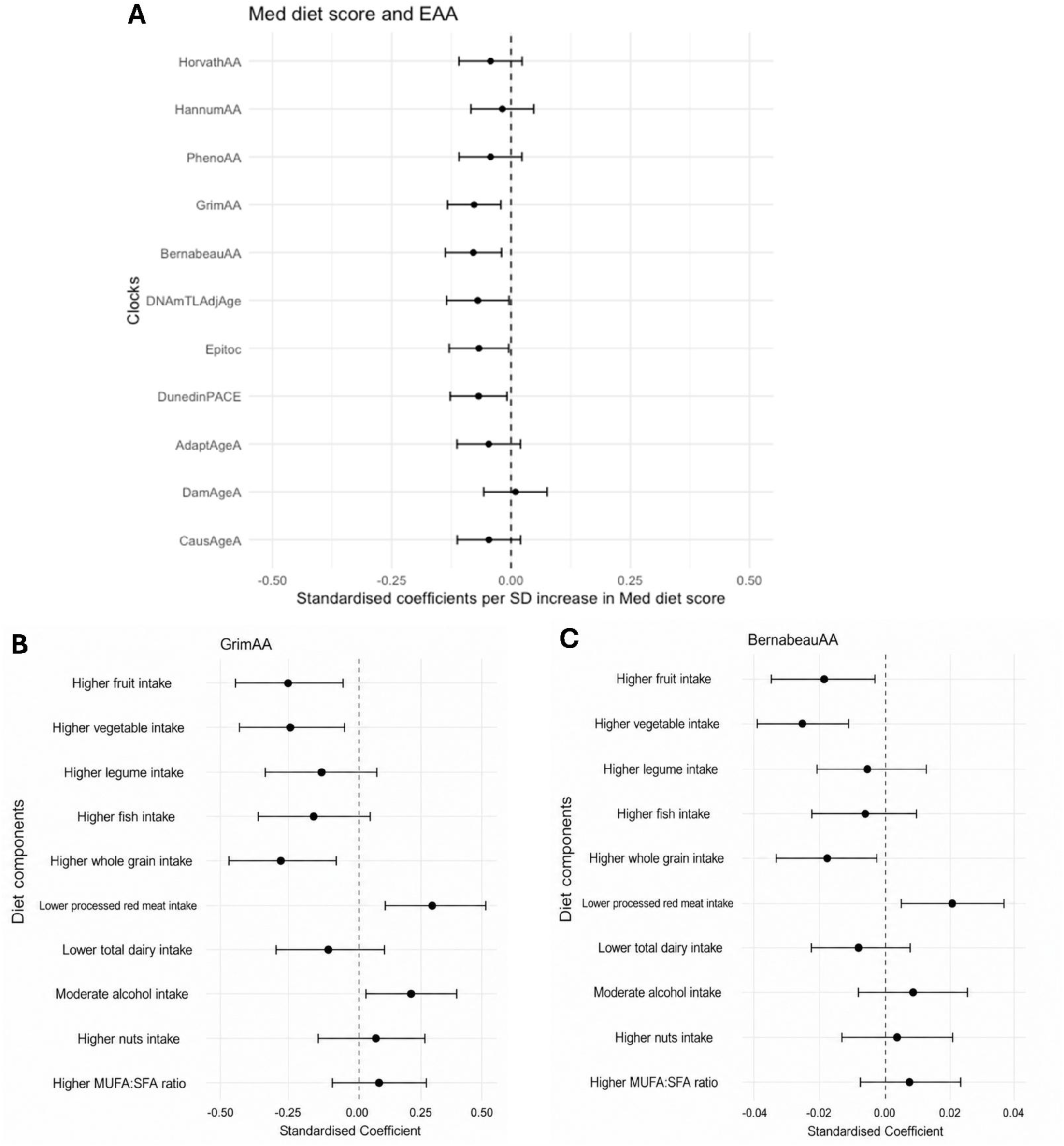
(**A**) Association between Mediterranean Diet Score (MDS) and EAA across 11 epigenetic clocks in 928 participants from the Airwave Health Monitoring Study. Effect estimates represent standardised beta coefficients per 1 SD increase in MDS, with 95% confidence intervals. Negative coefficients indicate lower EAA with greater adherence to the Mediterranean diet. (**B–C**) Associations between individual Mediterranean diet components and EAA as measured by GrimAge (**B**) and BernabeuAA (**C**). Effect estimates represent standardised beta coefficients with 95% confidence intervals. For all panels, models were adjusted for age, sex, BMI, household income, education level, alcohol intake, physical activity, anxiety, smoking status and total energy intake.

To identify dietary components contributing to these associations, we examined individual Mediterranean diet components in relation to GrimAge and BernabeuAA (**Fig. 1B–C, Table S3**). For GrimAge, higher fruit intake (β = −0.25 SD, 95% CI: −0.44, −0.05), vegetable intake (β = −0.24 SD, 95% CI: −0.43, −0.05), and whole grain intake (β = −0.27 SD, 95% CI: −0.46, −0.08) were associated with lower EAA, while lower processed red meat intake was associated with higher EAA (β = 0.27 SD, 95% CI: 0.07, 0.47); all remained significant following FDR correction. For BernabeuAA, higher vegetable intake was associated with lower EAA (β = −0.02 SD, 95% CI: −0.039, −0.011; FDR p = 0.005), whereas inverse associations for fruit and whole grain intake did not remain significant after multiple-testing correction.

### Smoking and EAA

In fully adjusted models (**Fig. 2A, Table S4**), current smokers had higher epigenetic age acceleration compared to never smokers for PhenoAge (0.27 SD; 95% CI: 0.06, 0.49; FDR p = 0.043), GrimAge (1.56 SD; 95% CI: 1.37, 1.74; FDR p < 0.001), BernabeuAA (1.41 SD; 95% CI: 1.22, 1.61; FDR p < 0.001), and DunedinPACE (0.69 SD; 95% CI: 0.50, 0.89; FDR p < 0.001). Among former smokers, higher EAA compared to never smokers was observed for GrimAge (0.50 SD; 95% CI: 0.37, 0.64; FDR p < 0.001), BernabeuAA (0.42 SD; 95% CI: 0.28, 0.56; FDR p < 0.001), and DunedinPACE (0.19 SD; 95% CI: 0.05, 0.33; FDR p = 0.025).

**Figure 2.**
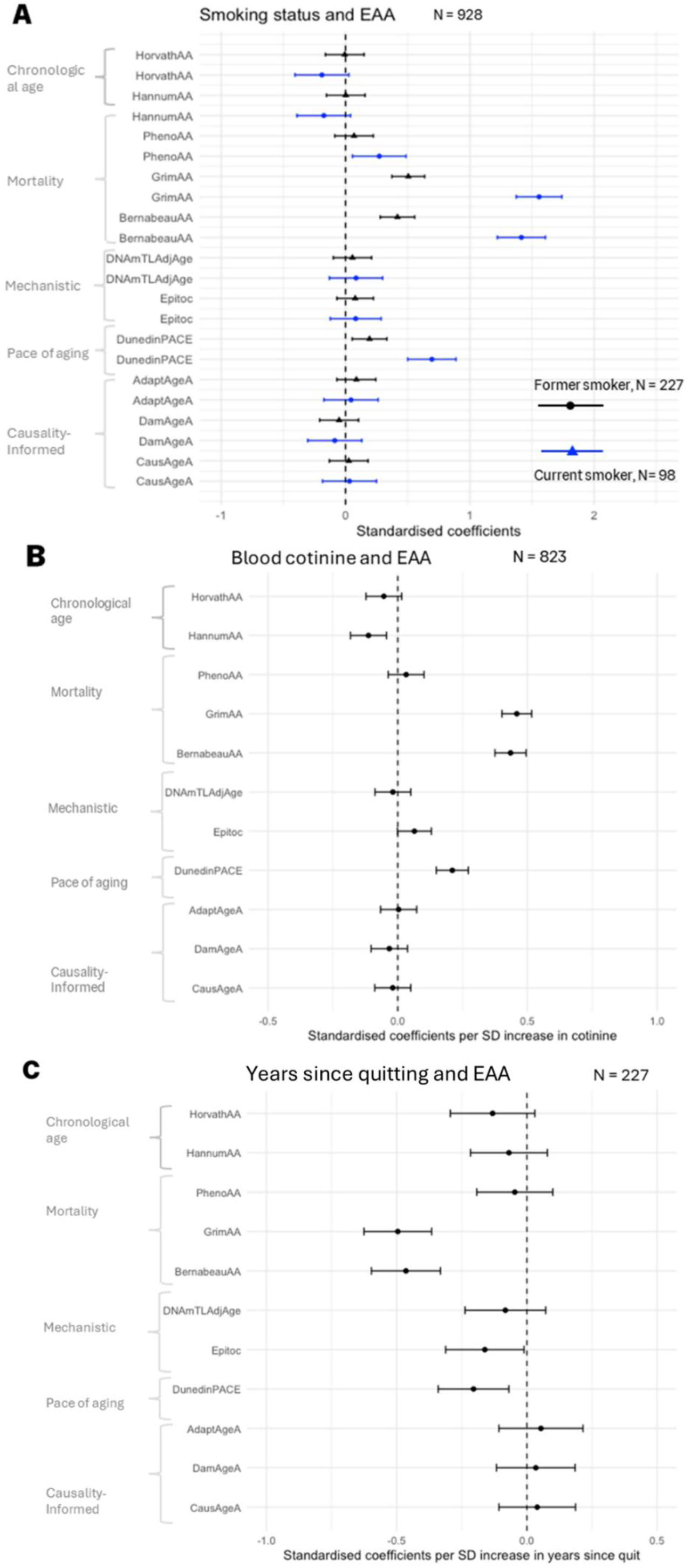
Fully adjusted regression models examining the association between smoking-related exposures and Epigenetic age acceleration (EAA). **A**: Comparison of current (n = 98) and former smokers (n = 227) to never smokers (ref: n = 603). **B**: Association between blood cotinine levels and EAA (n=823) and **C**: Association between time since quitting smoking (in years) and EAA among former smokers (n = 227). Models were adjusted for age, sex, BMI, household income, education level, alcohol intake, MDS physical activity, and anxiety.

In analyses using blood cotinine levels (**Fig. 2B, Table S4**), higher cotinine was associated with higher EAA for GrimAge (0.46 SD; 95% CI: 0.40, 0.52; FDR p < 0.001), BernabeuAA (0.43 SD; 95% CI: 0.37, 0.49; FDR p < 0.001), and DunedinPACE (0.21 SD; 95% CI: 0.15, 0.27; FDR p < 0.001). An inverse association was observed for HannumAA (–0.11 SD; 95% CI: –0.18, –0.04; FDR p = 0.004). The association with EpiTOC (0.06 SD; 95% CI: –0.00, 0.13) did not remain statistically significant after correction (FDR p = 0.120).

Among former smokers (**Fig. 2C**, **Table S4**), time since quitting was inversely associated with EAA for GrimAge (–0.50 SD; 95% CI: –0.63, –0.37; FDR p < 0.001), BernabeuAA (–0.46 SD; 95% CI: –0.60, –0.33; FDR p < 0.001), and DunedinPACE (–0.20 SD; 95% CI: –0.34, –0.07; FDR p = 0.012). The association with EpiTOC (–0.16 SD; 95% CI: –0.31, –0.01) did not remain significant after correction (FDR p = 0.099). No consistent associations were observed for the first-generation clocks (HorvathAA, HannumAA), DNAmTLAdjAge, or the causality-enriched clocks (AdaptAA, DamAA, CausAA) across smoking exposures.

### Modification of effect of smoking on EAA, by diet

Stratified analyses by high vs. low MDS revealed differences in associations between smoking status and EAA across subgroups (**Fig. 3 Table S5**). Among former smokers, GrimAge and Bernabeu remained significantly associated with elevated EAA across both adherence groups but were attenuated with higher adherence. GrimAge showed a 0.57 SD increase in the low group (95% CI: 0.38, 0.77), compared to 0.37 SD in the high group (95% CI: 0.15, 0.60). Bernabeu was 0.47 SD (95% CI: 0.27, 0.68) in the low group vs. 0.24 SD (95% CI: 0.00, 0.49) in the high group. DunedinPACE estimates were similar across diet strata among former smokers: 0.19 SD (95% CI: −0.04, 0.41) in the low adherence group and 0.19 SD (95% CI: −0.04, 0.43) in the high adherence group. Interaction terms were statistically significant for GrimAge and BernabeuAA (both *P*_interaction_ < 0.001), but not for DunedinPACE (*P*_interaction_ = 0.50). Among current smokers, those with low diet adherence exhibited greater EAA compared to never smokers, especially for GrimAge (1.79 SD, 95% CI: 1.54, 2.04) and Bernabeu (1.63 SD, 95% CI: 1.36, 1.90). These effects were attenuated in the high adherence group, with GrimAge at 1.35 SD (95% CI: 1.01, 1.68) and Bernabeu at 1.11 SD (95% CI: 0.74, 1.48). Interaction terms confirmed statistically significant moderation by MDS for GrimAge (P_interaction_ < 0.001), whereas evidence for interaction with BernabeuAA was weaker and did not reach statistical significance (*P*_interaction_ = 0.057). For DunedinPACE, effect estimates were also higher among individuals with lower diet adherence (0.78 SD; 95% CI: 0.49, 1.08) compared with those with higher adherence (0.56 SD; 95% CI: 0.21, 0.90); however, no interaction was observed for DunedinPACE (P_interaction_ = 0.14).

**Figure 3.**
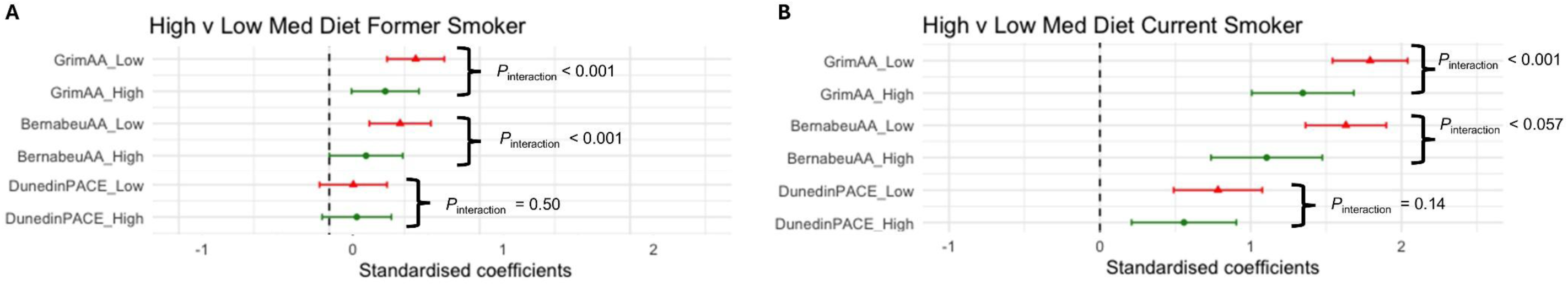
Standardised coefficients (β) with 95% confidence intervals from fully adjusted linear regression models showing the association between smoking status and epigenetic age acceleration (EAA), stratified by Mediterranean Diet Score (MDS; high vs. low, defined by cohort median). (**A**) Former vs. never smokers (n = 227 former smokers). (**B**) Current vs. never smokers (n = 98 current smokers). Models were adjusted for age, sex, BMI, household income, education, alcohol intake, physical activity, and anxiety. P_interaction values were derived from models including a continuous MDS × smoking status interaction term and are shown for selected clocks.

Further interaction analyses of individual Mediterranean diet components showed that several food groups modified the association between smoking and epigenetic age acceleration, particularly for GrimAge and BernabeuAA (**Fig. 4, Table S6**). For GrimAge, interactions terms for higher fruit intake (−0.12 SD, 95% CI: −0.19, −0.05), vegetables (−0.08 SD, 95% CI: −0.15, −0.01), whole grains (−0.12 SD, 95% CI: −0.19, −0.05), and lower dairy intake (0.09 SD, 95% CI: 0.02, 0.16) were significant after correction for multiple testing. For BernabeuAA, significant interactions were observed for higher vegetable intake (−0.12 SD, 95% CI: −0.19, −0.05) and higher whole grain intake (−0.11 SD, 95% CI: −0.18, −0.04). Associations for other components, including fruit and fish intake, were attenuated after correction for multiple testing.

**Figure 4.**
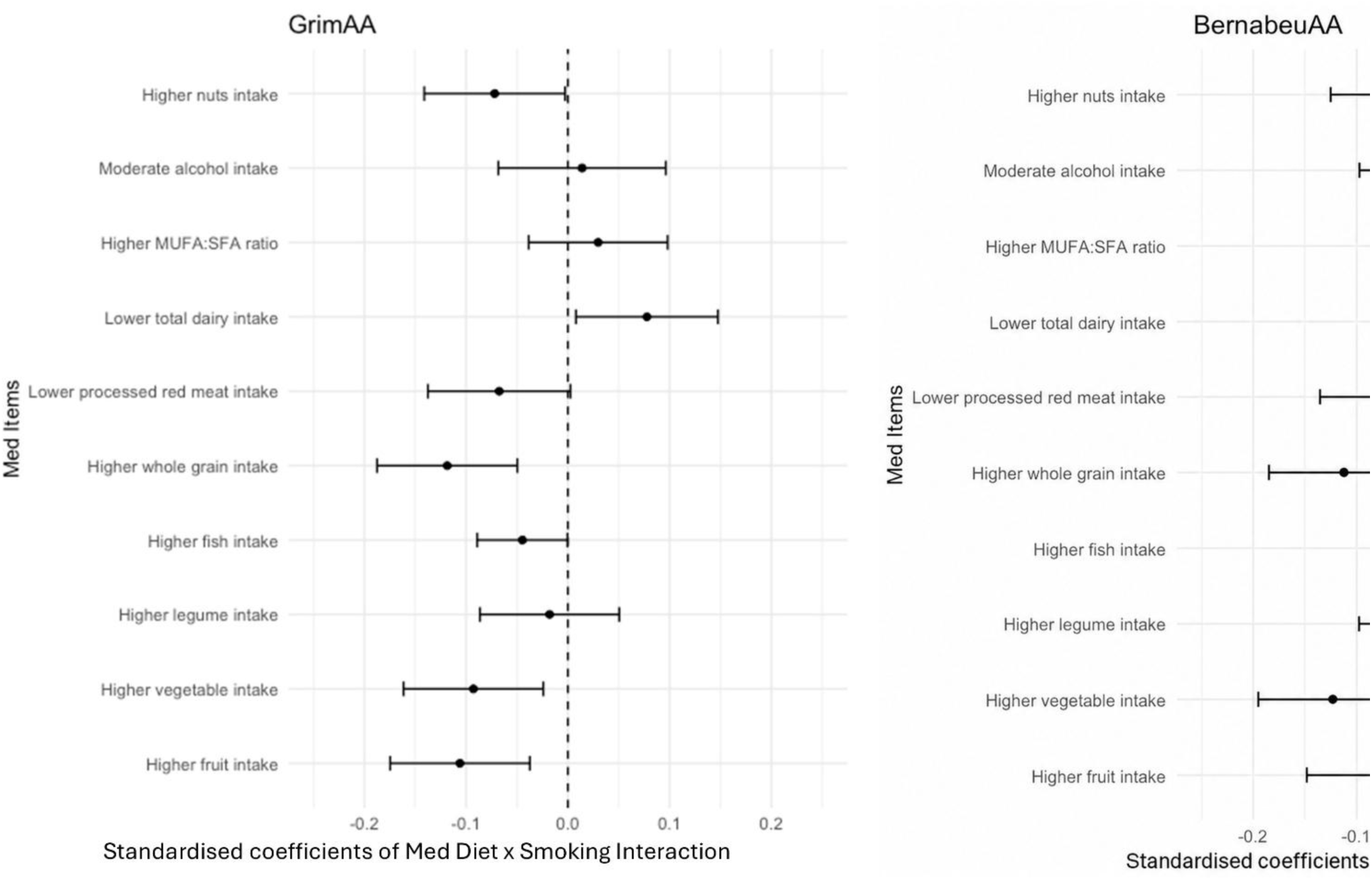
Standardised coefficients (with 95% CIs) from fully adjusted regression models showing the interaction effects between smoking status and individual Mediterranean Diet components on EAA, for GrimAge and BernabeuAA. Each point represents the standardised coefficient for the interaction term (Smoking × Diet component), indicating how dietary intake modifies the association between smoking and EAA. Negative values suggest that higher intake of the component attenuates smoking-related age acceleration. Models were adjusted for age, sex, BMI, household income, education level, alcohol intake, physical activity, anxiety and total energy intake.

### Sensitivity analysis

Sensitivity analyses comparing dietary under-reporters with plausible reporters showed no significant differences in GrimAge, BernabeuAA or DunedinPACE after adjustment for age, sex and BMI (**Table S7**).

## DISCUSSION

This study demonstrates that later-generation epigenetic clocks effectively capture the biological tension between harmful environmental stressors and protective lifestyle factors. We found that higher adherence to a Mediterranean diet was associated with significantly lower epigenetic age acceleration. This relationship was particularly second-generation mortality-trained clocks such as Bernabeu and GrimAge. By using smoking as a high-contrast benchmark, we further established that these specific clocks could detect the potential for dietary patterns to attenuate the molecular damage associated with tobacco smoke exposure.

Our findings were consistent across multiple clocks and reinforce growing evidence that higher Mediterranean diet adherence is associated with slower epigenetic ageing. For example, our results closely align with those of Kim and colleagues [20], who reported inverse associations between Mediterranean diet scores and both GrimAge and the precursor to DunedinPACE. This is further supported by recent findings from the Women’s Health Initiative, in which higher adherence to Healthy Eating Index, DASH, and alternative Mediterranean dietary patterns was associated with lower epigenetic age acceleration, particularly for GrimAge, PhenoAge, and DunedinPACE across 4,500 postmenopausal women [52]. Consistent with our findings, associations were strongest for second-generation ageing biomarkers and measures of pace of ageing. We observed comparable effect sizes of approximately –0.08 standard deviations per 1 standard deviation increase in diet score. Further supporting evidence from the NU-AGE intervention study showed that a one-year Mediterranean-style dietary intervention was associated with lower Horvath-derived epigenetic age acceleration measures [53]. We extend these findings to additional clocks, including Bernabeu, EpiTOC, and DNAmTL, although we did not replicate the PhenoAge association reported in previous studies.

The strong associations observed for smoking provide an internal benchmark for interpreting these findings. Current smokers exhibited pronounced age acceleration relative to non-smokers, with GrimAge showing the strongest response. Prior studies have associated smoking with a reduction in life expectancy of ∼10 years [31]. Given that a 1-year GrimAge acceleration approximately corresponds to a hazard ratio of 1.10 for mortality [7], and we observed a 7.18-year acceleration among current smokers, this implies nearly a doubling of mortality risk (HR = 1.95), aligning well with existing evidence of significant life expectancy reductions in smokers [54]. This finding is expected, given that Grimage, like Benabeu, incorporated smoking history into its training algorithm. PhenoAge is driven by a limited selection of broad systemic markers like inflammation and metabolic dysfunction, thus smoking’s impact may be diluted, especially in those with healthier baseline physiology or former smokers. While PhenoAge captures long-term aging risk, it appears less sensitive to smoking. In contrast, DunedinPACE more effectively captured smoking-related aging.

DunedinPACE incorporates markers across multiple systems such as cardiovascular (e.g., mean arterial pressure), pulmonary (FEV1), metabolic (BMI, HbA1c) and immune (CRP), all known to be impacted by tobacco exposure [11]. Our finding that GrimAge and DunedinPACE were most sensitive to smoking mirrors the ranking reported by Klopack et al. (2022) [55], who assessed lifetime smoking exposure, reinforcing the relative sensitivity of these clocks to tobacco exposure.

Blood cotinine levels further reinforced these trends, as dose-dependent associations with GrimAge, Bernabeu, and DunedinPACE. EpiTOC, a mitotic clock capturing cell division rates, showed a borderline association with cotinine, suggesting that smoking-induced replication stress contributes to accelerated aging. Additionally, consistent with prior tissue-based findings [56], our blood-based analysis shows that longer time since smoking cessation is associated with reduced epigenetic age acceleration as measured by in GrimAge, Bernabeu, DunedinPACE, with a nominal association observed for EpiTOC. On the contrary, some clocks, particularly the first-generation Horvath and Hannum clocks, as well as the causality-enriched clocks (AdaptAge, CausAge, and DamAge), did not demonstrate significant associations with smoking indices. Horvath and Hannum clocks, designed to predict chronological age, showed no significant associations with smoking, suggesting their reliance on time-dependent CpG changes rather than those influenced by external stressors.

Given the consistent diet associations across clocks, we examined whether diet modifies susceptibility to smoking-related age acceleration. For the clocks most associated with smoking, including DunedinPACE, GrimAge, and Bernabeu, the associations with smoking status were generally stronger among those with low Mediterranean diet adherence. Since these clocks partially reflect the methylation profile of smoking, this observation cannot be interpreted as an effect on biological ageing specifically. It does, however, imply that the mortality-related epigenetic imprinting of smoking is lessened by adhering to a nutritious diet. This interaction was most evident in GrimAge, where the antioxidant and anti-inflammatory properties of a Mediterranean pattern appear to buffer the oxidative engine of tobacco exposure. Component-level analyses indicated that higher intake of fruits, vegetables, and whole grains contributed most strongly to this attenuation, consistent with broader evidence linking these dietary patterns to reduced systemic inflammation and oxidative stress [57–59]. These findings closely mirror those reported by Kim et al. [19], who observed inverse associations between fruit, vegetable, whole grain, and legume intake and GrimAge acceleration. The consistency of these component-level associations across cohorts suggests that specific features of high-quality dietary patterns may be important determinants of biological ageing. Notably, Kim et al. also reported stronger associations between dietary components and epigenetic ageing among ever-smokers than never-smokers, which is consistent with our observation that dietary patterns appear most informative in the context of smoking-related ageing signals.

Interestingly, the observation that fibre-rich dietary components are associated with modification of smoking-related age acceleration is supported by a recent study showing that higher fibre intake was significantly associated with greater odds of healthy ageing in a large cohort [24]. These findings are also biologically plausible given experimental evidence that a polyphenol-rich Mediterranean dietary intervention can induce widespread changes in DNA methylation and gene expression, particularly in pathways related to inflammation and one-carbon metabolism [60]. These lines of evidence suggest that a healthy diet may buffer the biological impact of environmental pollutants more generally. Such results may be applicable to other oxidative stressors, such as ambient air pollution, which operate through similar pathways of inflammation and metabolic disruption.

### Strengths and Limitations

This study provides a comprehensive evaluation of the sensitivity of epigenetic clocks to smoking and examines the moderating influence of diet. Midlife represents a pivotal period in which lifestyle exposures exert measurable effects on biological ageing. In such populations, where extensive longitudinal health data or overt disease may be absent, epigenetic clocks provide a distinct advantage for quantifying biological impact. The large cohort size and the use of detailed smoking measures, including both self-reported status and blood cotinine, enhance the robustness of our findings. Furthermore, the use of seven-day food diaries allows for a more detailed and accurate assessment of dietary intake compared to food frequency questionnaires or 24-hour recalls. This enabled an in-depth analysis of specific Mediterranean diet components.

Several limitations must be considered. The cross-sectional design limits the ability to draw causal inferences, and the predominantly white occupational cohort may restrict the generalisability of the findings. A key limitation is the distribution of smoking status, with approximately six times more non-smokers than smokers. This imbalance reflects the contemporary decline in smoking prevalence and the occupational nature of the cohort, but it also introduces a potential healthy worker effect that may lead to an underestimation of smoking-related age acceleration. The relatively small number of current smokers with high Mediterranean diet adherence reduces statistical power, meaning results in these subgroups should be interpreted with caution. Additionally, participants who completed food diaries may differ from those who did not, introducing potential selection bias. In line with previous studies [61], we noted a proportion of participants were classified as under-reporters using the Goldberg method; however, sensitivity analyses indicated that dietary misreporting is unlikely to materially explain the observed associations. Furthermore, models examining Mediterranean diet exposures were adjusted for total energy intake, reducing the potential impact of systematic under-reporting on the observed associations. Future studies should incorporate objective dietary validation methods to minimise self-reporting bias. These might include metabolomic profiling or the measurement of specific nutrients.

## Conclusion

In conclusion, this study provides a critical evaluation of diverse epigenetic ageing biomarkers by examining their responsiveness to, and the interactions of, healthy diet and smoking, key protective and hazardous components of the exposome. The observed impact of specific dietary components, particularly fruits, vegetables, and whole grains, on smoking-related age acceleration reinforces the biological plausibility of these clocks. Specifically, the attenuation of age acceleration in clocks incorporating smoking-related scores suggests that methylation changes are not merely static markers of exposure. Instead, they reflect dynamic biological processes that can be influenced by lifestyle factors. This finding serves as a compelling evaluation of the ability of second-generation clocks to capture biologically meaningful and modifiable aspects of ageing. It is reasonable to postulate that the increased risk of age-related disease associated with smoking may be exacerbated by the absence of a healthy diet, which underscores the potential of dietary interventions to influence biological resilience.

## Data Availability

Data may be made available to qualified researchers upon reasonable request and subject to approval by the AIRWAVE data access committee and relevant governance procedures.

## Abbreviations

AIRWAVE: Airwave Health Monitoring Study
BMI: Body Mass Index
CI: Confidence Interval
CpG: Cytosine-phosphate-Guanine site
CRP: C-Reactive Protein
DNAm: DNA Methylation
DNAmTLAdjAge: DNA Methylation-Based Telomere Length Adjusted Age
EAA: Epigenetic Age Acceleration
HbA1c: Glycated Haemoglobin
IL-6: Interleukin-6
IQR: Interquartile Range
LTL: Leukocyte Telomere Length
MDS: Mediterranean Diet Score
SD: Standard Deviatio
SES: Socioeconomic Status
TL: Telomere Length.

## AUTHOR CONTRIBUTIONS

IM, OR and AZ conceived the study. AZ performed analyses and drafted the manuscript. CL prepared and supervised data collection in AIRWAVE cohort. IM, OR, CL, RE and GF revised manuscript. All authors critically reviewed manuscript.

## ACKNOWLEDGEMENTS

AZ was supported by the Medical Research Council Centre for Environment and Health (MR/S019669/1), UKRI Dream (NE/T001895/1). The Airwave Health Monitoring Study is funded by the Home Office (grant number 780-TETRA) with additional support from the National Institute for Health Research (NIHR) Biomedical Research Centre. The Airwave Study uses the computing resources of the UK MEDical BIOinformatics Partnership (UK MED-BIO: supported by the Medical Research Council MR/L01632X/1). OR and CHL were supported by a UKRI Future Leaders Fellowship (MR/S03532X/1, MR/Y02012X/1). We thank all Airwave participants for their contributions.

## DATA SHARING

This research has been conducted with the AIRWAVE Health and Monitoring Study. Data are available upon request.

## ETHICAL STATEMENT

Ethical approval was obtained from the National Health Service Multi-Site Research Ethics Committee (MREC/13/NW/0588). All participants provided written informed consent.

## Supporting Information

Table S1: Summary of DNA methylation ageing clocks. Tables S2–S6: Additional regression analyses of Mediterranean diet, smoking, and their interactions with epigenetic age acceleration. Table S7: Sensitivity analysis of Goldberg-defined dietary under-reporting. Figures S1–S2: Study design and directed acyclic graph. Figures S3–S5: Validation and descriptive analyses of smoking, Mediterranean diet, and epigenetic clock performance.

