## Supplementary Material for "The role of Mediterranean diet adherence, smoking and their interactions in epigenetic age acceleration: A cross-sectional analysis of the Airwave cohort"

### **SUPPORTING INFORMATION**

Table S1 - A summary of various DNAm clocks, highlighting their key characteristics such as the number of CpGs, applicable age ranges, sample sizes, tissues, and development platforms.

| Epigenetic Clock | Training Population | Platform | Number of CpGs Used | Age Range | Sample size | Tissues Used | Trained on | Training Method | Biomarkers Used |
| --- | --- | --- | --- | --- | --- | --- | --- | --- | --- |
| Horvath | Multiple tissues from assorted studies | Illumina 27K & 450K | 353 | 0-101 | 8000 | Multi-tissue | Chronological age | Trained using elastic net regression on DNAm data across multiple tissues |  |
| Hannum Clock | Whole blood samples from Caucasian (64.9%) and Hispanic (35.1%) individuals, | Illumina 450K | 71 | 19-101 | 656 | Blood | Chronological age | Trained using linear regression on DNAm data from blood samples |  |
| PhenoAge | NHANES IV participants, ~ 75% Non-Hispanic White, Non-Hispanic Black ~ 15%, Mexican American ~ 10%, USA | Illumina 27K,  450K & EPIC | 513 | 20-85 | 9926 | Blood | Phenotypic age (panel of clinical markers, selected on mortality) | Trained using elastic net regression on phenotypic age derived from clinical markers and DNAm data | Albumin, Creat, Glu, CRP, Lymph%, MCV, RDW, ALP, WBC |
| GrimAge | Framingham Heart Study, 100% European American, Massachusetts, USA | Illumina 450K & EPIC | 1030 | 40-92 | 1731 | Blood | Two-step process including proteins and pack years trained on Mortality risk | Trained using elastic net regression to predict lifespan based on DNAm + plasma proteins (e.g., smoking pack-years) | Adrenomedullin, Beta-2-Microglobulin, Cystatin C, Growth Differentiation Factor 15, Leptin, Plasminogen Activator Inhibitor-1, Tissue Inhibitor Metalloproteinases 1, Smoking Pack-Years |
| Bernabeu | Generation Scotland cohort and external cohorts | Illumina EPIC array | 2330 | 18-99 | 18,365 | Blood | time to all-cause mortality | Elastic net regression with feature pre-selection and LOCO framework | GrimAge components (smoking pack years, seven plasma proteins) and twenty-eight protein EpiScores described in (Gadd et al., 2022) |
| DunedinPACE | Dunedin Study cohort, 92.5% European descent, 7.5% Maori | Illumina 450K | 46 | 26-38 (repeated measures) | 1037 | Blood | Pace of ageing (change over three timepoint of clinical markers) | Trained using elastic net regression to track the pace of ageing across 20 years | BMI, WHR, HbA1c, Leptin, MAP, VO2Max, FEV1/FVC, FEV1, TC, TG, HDL, Lp(a), ApoB/A1, eGFR, BUN, hs-CRP, WBC, AL, Caries |
| DNAmTL | Women’s Health Initiative and Jackson Heart Study cohort, Female: 72% (100% in WHI, 64% in JHS), European Ancestry (EUR): 20.2% (59% in WHI)  African Ancestry (AFR): 79.8% (41% in WHI, 100% in JHS) | Illumina 450K & EPIC | 140 | 22-93 | 2256 | Blood | Telomere length | Trained to estimate telomere length based on DNAm data |  |
| Epitoc | Healthy individuals with data from normal and cancerous tissues | Illumina 450K & EPIC | 385 PCGT/PRC2-marked promoter CpGs | Wide age range includes ages from fetal tissue to 80+ years) | 656 whole blood samples from Hannum dataset | Whole blood, purified cell subtypes, various normal and cancerous tissues, buccal tissue, stem cell populations | cell division rates | Linear regression of age versus DNA methylation, adjusted for cell type composition, followed by validation in various datasets | PCGT (Polycomb group target) promoter CpGs, PRC2-marked promoter CpGs |
| AdaptAge | Generation Scotland | Illumina 450K | 1000 | Various | 2664 | Blood, multiple tissues | CpG sites with protective effects, both identified by causal effect size and direction of age-associated differential methylation | Two-sample Mendelian Randomization |  |
| DamAge | Generation Scotland | Illumina 450K | 1090 | Various | 2664 | Blood, multiple tissues | CpG sites with damaging effects. | Two-sample Mendelian Randomization |  |
| CausAge | Generation Scotland | Illumina 450K | 586 | Various | 2664 | Blood, multiple tissues | CpG sites identified as causal to adjusted Aging-GIP1 | Two-sample Mendelian Randomization |  |

Note: Adjusted Aging-GIP1 is a genetic principal component that captures the shared genetic basis of six ageing-related traits: health span, parental lifespans, exceptional longevity, frailty index, and self-rated health, adjusted for socioeconomic factors.

*Table S2* - Fully adjusted model showing associations between Mediterranean Diet score and EAA across various clocks. Standardized coefficients (95% confidence intervals) per standard deviation increase in Mediterranean diet Score. Models were adjusted for age, sex, BMI, household income, education level, alcohol intake, physical activity, anxiety, smoking status and total energy intake.

| **Clock** | **Coef. 95CI** | **P-value** |
| --- | --- | --- |
| HorvathAA | -0.04 (-0.11, 0.03) | 0.192 |
| HannumAA | -0.02 (-0.09, 0.05) | 0.585 |
| PhenoAA | -0.04 (-0.11, 0.03) | 0.211 |
| GrimAA | -0.07 (-0.13, -0.01) | 0.004* |
| BernabeuAA | -0.08 (-0.02, -0.14) | 0.006* |
| DNAmTLAdjAge | -0.06 (-0.13, 0.00) | 0.036 |
| Epitoc | -0.07 (-0.00, -0.13) | 0.033 |
| DunedinPACE | -0.07 (-0.01, -0.13) | 0.025 |
| AdaptAA | -0.04 (-0.10, 0.03) | 0.171 |
| DamAA | 0.00 (-0.06, 0.07) | 0.759 |
| CausAA | -0.04 (-0.11, 0.03) | 0.170 |

- Coefficients represent standardized effect sizes (β) with 95% confidence intervals (CI) for the association between Mediterranean Diet Score and EAA. P-values indicate the statistical significance of these associations.
- * Indicates associations that remain statistically significant after correction for multiple testing using Benjamini–Hochberg FDR applied across all tests

Table S3 - Fully adjusted model showing associations between individual Mediterranean diet components and epigenetic age acceleration as measured by GrimAge and BernabeuAA. Standardized coefficients (95% confidence intervals) per unit increase in Mediterranean diet component score. Models were adjusted for age, sex, BMI, household income, education level, alcohol intake, physical activity, anxiety, smoking status and total energy intake.

| **Dietary Component** | **GrimAA Coef. (95% CI)** | **P-value** | **BernabeuAA Coef. (95% CI)** | **P-value** |
| --- | --- | --- | --- | --- |
| Higher fruit intake | -0.25 (-0.44, -0.05) | 0.013* | -0.02 (-0.03, -0.00) | 0.018 |
| Higher vegetable intake | -0.24 (-0.43, -0.05) | 0.013* | -0.02 (-0.04, -0.01) | <0.001* |
| Higher legume intake | -0.14 (-0.33, 0.05) | 0.159 | -0.01 (-0.02, 0.01) | 0.362 |
| Higher fish intake | -0.16 (-0.35, 0.03) | 0.096 | -0.01 (-0.02, 0.01) | 0.273 |
| Higher whole grain intake | -0.27 (-0.46, -0.08) | 0.006* | -0.02 (-0.03, -0.00) | 0.018 |
| Lower processed red meat intake | 0.27 (0.07, 0.47) | 0.008* | 0.02 (0.00, 0.03) | 0.034 |
| Lower total dairy intake | -0.10 (-0.30, 0.09) | 0.300 | -0.01 (-0.02, 0.01) | 0.318 |
| Moderate alcohol intake | 0.23 (0.03, 0.43) | 0.028 | 0.01 (-0.01, 0.02) | 0.351 |
| Higher nuts intake | 0.06 (-0.13, 0.25) | 0.523 | 0.00 (-0.01, 0.02) | 0.665 |
| Higher MUFA:SFA ratio | 0.09 (-0.10, 0.28) | 0.364 | 0.01 (-0.01, 0.02) | 0.401 |

- Coefficients represent standardized effect sizes (β) with 95% confidence intervals (CI) for the association between individual Mediterranean diet components and epigenetic age acceleration (EAA).
- P-values indicate the statistical significance of these associations.
- * Indicates associations that remain statistically significant after correction for multiple testing using the Benjamini–Hochberg false discovery rate (FDR) applied separately across all dietary component tests for each clock.

*Table S4* - Fully adjusted model for current (n =98) and former smokers (n=227), compared to never smokers (n=603). Estimates show difference in scaled EAA with 95% confidence intervals. Adjusted for age, sex, household income, education level, physical activity, Mediterranean Diet score, BMI, alcohol consumption and anxiety levels. Standardized coefficients with 95% confidence intervals (CI) for smoking status, blood cotinine (n=823), and time since quitting (n=227) across epigenetic clocks.

| **Clock** | **Smoking status (current vs. never)** | **P-value** | **Smoking status (former vs. never)** | **P-value** | **Blood cotinine (n = 823)** | **P-value** | **Time since quitting (former smokers)** | **P-value** |
| --- | --- | --- | --- | --- | --- | --- | --- | --- |
| HorvathAA | -0.19 (-0.41, 0.03) | 0.101 | -0.03 (-0.19, 0.13) | 0.624 | -0.05 (-0.12, 0.02) | 0.125 | -0.11 (-0.29, 0.06) | 0.111 |
| HannumAA | -0.17 (-0.39, 0.04) | 0.102 | 0.00 (-0.16, 0.16) | 0.922 | -0.11 (-0.18, -0.04) | 0.001* | -0.05 (-0.21, 0.11) | 0.360 |
| PhenoAA | 0.27 (0.06, 0.49) | 0.016 | 0.09 (-0.07, 0.26) | 0.236 | 0.03 (-0.04, 0.10) | 0.359 | -0.05 (-0.20, 0.11) | 0.533 |
| GrimAA | 1.56 (1.37, 1.74) | <0.001* | 0.50 (0.37, 0.64) | <0.001* | 0.46 (0.40, 0.52) | <0.001* | -0.50 (-0.63, -0.37) | <0.001* |
| BernabeuAA | 1.41 (1.22, 1.61) | <0.001* | 0.42 (0.28, 0.56) | <0.001* | 0.43 (0.37, 0.49) | <0.001* | -0.46 (-0.60, -0.33) | <0.001* |
| DNAmTLAdjAge | 0.08 (-0.14, 0.29) | 0.440 | 0.05 (-0.11, 0.21) | 0.302 | -0.02 (-0.09, 0.05) | 0.591 | -0.08 (-0.25, 0.09) | 0.295 |
| Epitoc | 0.09 (-0.12, 0.29) | 0.435 | 0.07 (-0.08, 0.22) | 0.300 | 0.06 (-0.00, 0.13) | 0.055 | -0.16 (-0.31, -0.01) | 0.036* |
| DunedinPACE | 0.69 (0.50, 0.89) | <0.001* | 0.19 (0.05, 0.33) | 0.007* | 0.21 (0.15, 0.27) | <0.001* | -0.20 (-0.34, -0.07) | 0.003* |
| AdaptAgeA | 0.05 (-0.17, 0.27) | 0.693 | 0.09 (-0.07, 0.25) | 0.516 | 0.00 (-0.07, 0.07) | 0.924 | 0.05 (-0.12, 0.22) | 0.510 |
| DamAgeA | -0.09 (-0.31, 0.13) | 0.433 | -0.03 (-0.19, 0.13) | 0.748 | -0.03 (-0.10, 0.04) | 0.363 | -0.01 (-0.17, 0.16) | 0.655 |
| CausAgeA | 0.03 (-0.19, 0.25) | 0.775 | 0.03 (-0.14, 0.19) | 0.085 | -0.02 (-0.09, 0.05) | 0.583 | 0.06 (-0.09, 0.21) | 0.595 |

- Multiple testing correction was performed using the Benjamini–Hochberg false discovery rate (FDR). Coefficients
- represent standardized effect sizes (β) with 95% confidence intervals (CI). Each column shows the association between smoking-related variables and EAA
- * Indicates associations that remain statistically significant after correction for multiple testing using Benjamini–Hochberg FDR applied across all tests.
- P-values indicate the statistical significance of these associations.

Table S5 - Fully adjusted model for current smokers compared to never smokers, stratified by high (above median) or low (below median) Mediterranean diet, and EAA (Standardized Coefficients with 95% Confidence Intervals). Adjusted for age, sex, BMI, income, education, alcohol consumption, physical activity, anxiety, and total energy intake

| **Clock** | **Former Smokers (High Med Diet)** | **P-value** | **Former Smokers (Low Med Diet)** | **P-value** | | **Current Smokers (High Med Diet)** | **P-value** | **Current Smokers (Low Med Diet)** | **P-value** |
| --- | --- | --- | --- | --- | --- | --- | --- | --- | --- |
| HorvathAA | -0.04 (-0.32, 0.24) | 0.259 | 0.05 (-0.19, 0.30) | | 0.463 | -0.06 (-0.46, 0.35) | 0.164 | -0.17 (-0.50, 0.15) | 0.386 |
| HannumAA | 0.02 (-0.26, 0.31) | 0.675 | -0.07 (-0.31, 0.17) | | 0.699 | 0.13 (-0.29, 0.55) | 0.161 | -0.14 (-0.43, 0.15) | 0.253 |
| PhenoAA | 0.03 (-0.24, 0.30) | 0.819 | -0.07 (-0.30, 0.17) | | 0.504 | 0.37 (-0.04, 0.79) | 0.075 | 0.25 (-0.06, 0.56) | 0.213 |
| GrimAA | 0.37 (0.15, 0.60) † | <0.001* | 0.57 (0.38, 0.77) † | | <0.001* | 1.35 (1.01, 1.68) † | <0.001* | 1.79 (1.54, 2.04) † | <0.001 |
| BernabeuAA | 0.24 (0.00, 0.49) ‡ | 0.052 | 0.47 (0.27, 0.68) ‡ | | <0.001* | 1.11 (0.74, 1.48) ‡ | <0.001 | 1.63 (1.36, 1.90) ‡ | <0.001 |
| DNAmTLAdjAge | 0.19 (-0.09, 0.47) | 0.313 | -0.02 (-0.26, 0.21) | | 0.926 | 0.18 (-0.23, 0.59) | 0.486 | -0.08 (-0.39, 0.23) | 0.272 |
| Epitoc | 0.10 (-0.15, 0.35) | 0.577 | 0.05 (-0.18, 0.28) | | 0.837 | 0.17 (-0.20, 0.53) | 0.796 | 0.06 (-0.25, 0.37) | 0.506 |
| DunedinPACE | 0.19 (-0.04, 0.43) | 0.118 | 0.19 (-0.04, 0.41) | | 0.161 | 0.56 (0.21, 0.90) | 0.002* | 0.78 (0.49, 1.08) | <0.001 |
| AdaptAgeA | 0.27 (-0.02, 0.56) | 0.683 | -0.04 (-0.27, 0.19) | | 0.726 | -0.06 (-0.48, 0.37) | 0.510 | -0.01 (-0.32, 0.31) | 0.834 |
| DamAgeA | -0.20 (-0.50, 0.10) | 0.513 | 0.07 (-0.15, 0.29) | | 0.945 | -0.04 (-0.48, 0.40) | 0.444 | -0.01 (-0.30, 0.29) | 0.776 |
| CausAgeA | 0.11 (-0.15, 0.37) | 0.116 | -0.02 (-0.26, 0.22) | | 0.334 | 0.07 (-0.31, 0.45) | 0.886 | -0.01 (-0.34, 0.31) | 0.970 |

- Coefficients represent standardized effect sizes (β) with 95% confidence intervals (CI), indicating the change in epigenetic age acceleration (EAA) per unit change in smoking status, stratified by Mediterranean Diet adherence (Low vs. High).
- † P_interaction = 0.001 for GrimAge in current smokers and <0.001 in former smokers.
- ‡ P_interaction = 0.057 for BernabeuAA in current smokers and <0.001 in former smokers.
- § P_interaction = 0.14 for DunedinPACE in current smokers and 0.50 in former smokers.
- P-values indicate the statistical significance of associations within strata.
- * Indicates associations that remain statistically significant after correction for multiple testing using Benjamini–Hochberg FDR applied across all tests.

Table S6 - Interaction effects of Mediterranean diet components on smoking-related epigenetic age acceleration as measured by GrimAge and Bernabeu clocks. Standardized Coefficients with 95% Confidence Intervals. Models were adjusted for age, sex, BMI, household income, education level, alcohol intake, physical activity, anxiety, smoking status and total energy intake.

| **Dietary Component** | **GrimAge (Coef. 95% CI)** | **P-value** | **Bernabeu (Coef. 95% CI)** | **P-value** |
| --- | --- | --- | --- | --- |
| Higher fruit intake | –0.12 (–0.19, –0.05) | 0.001* | –0.08 (–0.15, –0.00) | 0.040 |
| Higher vegetable intake | –0.08 (–0.15, –0.01) | 0.019 | –0.12 (–0.19, –0.05) | 0.001* |
| Higher legume intake | –0.01 (–0.08, 0.06) | 0.826 | –0.03 (–0.10, 0.05) | 0.486 |
| Higher fish intake | –0.05 (–0.09, –0.00) | 0.033 | –0.02 (–0.07, 0.02) | 0.297 |
| Higher whole grain intake | –0.12 (–0.19, –0.05) | 0.001* | –0.11 (–0.18, –0.04) | 0.003* |
| Lower processed red meat intake | –0.07 (–0.14, 0.00) | 0.065 | –0.06 (–0.14, 0.01) | 0.101 |
| Lower total dairy intake | 0.09 (0.02, 0.16) | 0.016 | 0.08 (0.00, 0.15) | 0.043 |
| Higher MUFA:SFA ratio | 0.04 (–0.03, 0.11) | 0.242 | 0.01 (–0.06, 0.08) | 0.820 |
| Moderate alcohol intake | 0.02 (–0.07, 0.10) | 0.708 | –0.01 (–0.10, 0.08) | 0.809 |
| Higher nuts intake | –0.06 (–0.13, 0.01) | 0.093 | –0.05 (–0.13, 0.02) | 0.161 |

- Coefficients represent standardized interaction effects (β) for each Mediterranean Diet component × smoking status on EAA. Negative coefficients indicate attenuation (reduction) of smoking-dependent age acceleration; positive coefficients suggest an enhancement.
- P-values indicate statistical significance of the interaction term. P < 0.05 is considered statistically significant.
- * Indicates interaction effects that remain statistically significant after correction for multiple testing using Benjamini–Hochberg FDR applied across all tests.

Table S7 - Association between Goldberg-defined under-reporting status and selected epigenetic ageing measures.

| **Outcome** | **Coef. (95% CI)** | **P-value** |
| --- | --- | --- |
| GrimAge | 0.11 (-0.02, 0.24) | 0.101 |
| BernabeuAA | 0.10 (-0.03, 0.24) | 0.126 |
| DunedinPACE | 0.06 (-0.06, 0.18) | 0.355 |

- Under-reporters (n = 415) were compared with plausible reporters (n = 513) according to the Goldberg method.
- Coefficients represent the difference in standardised epigenetic age acceleration between under-reporters and plausible reporters.
- Models were adjusted for age, sex, and BMI.
- Positive coefficients indicate higher epigenetic age acceleration among under-reporters relative to plausible reporters.

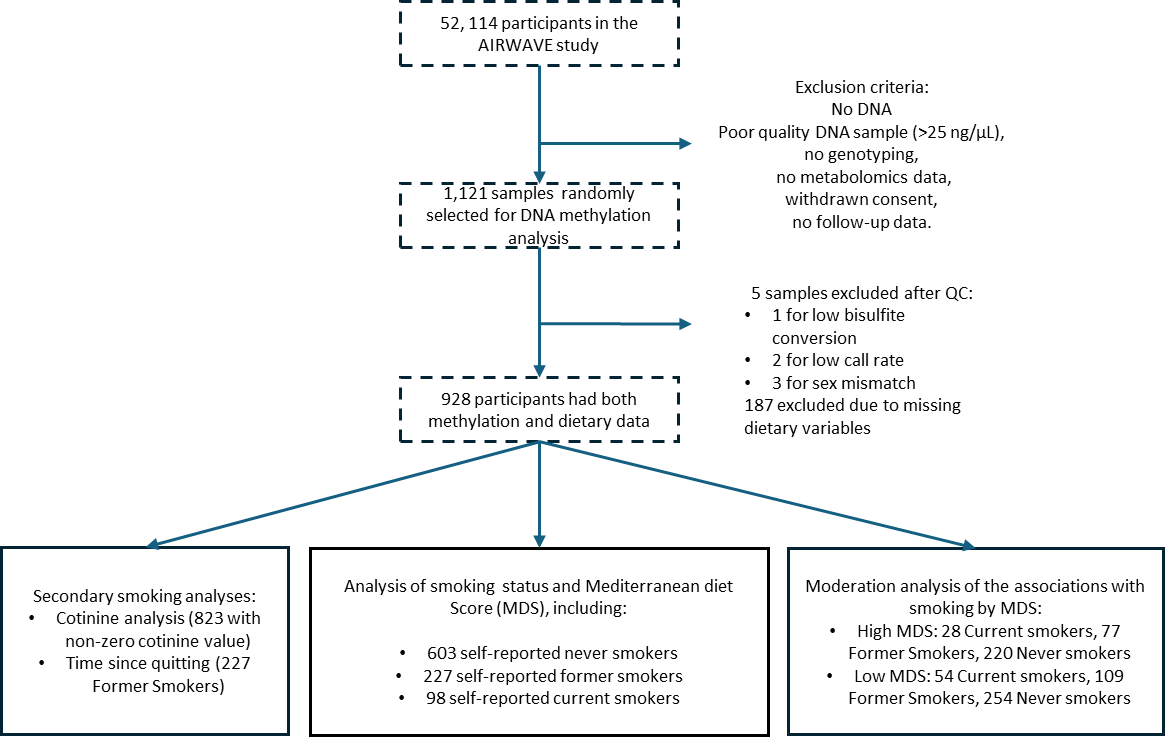

*Figure S1* Depiction of the study design and analyses. Participants were stratified by smoking status (current, former, and never smokers). Multivariate linear regression analysis was conducted to examine the relationship between smoking status and EAA, with further interaction analyses exploring the moderation of the Mediterranean diet. Secondary analyses were performed to evaluate the effects of blood cotinine levels and time since quitting smoking on EAA.

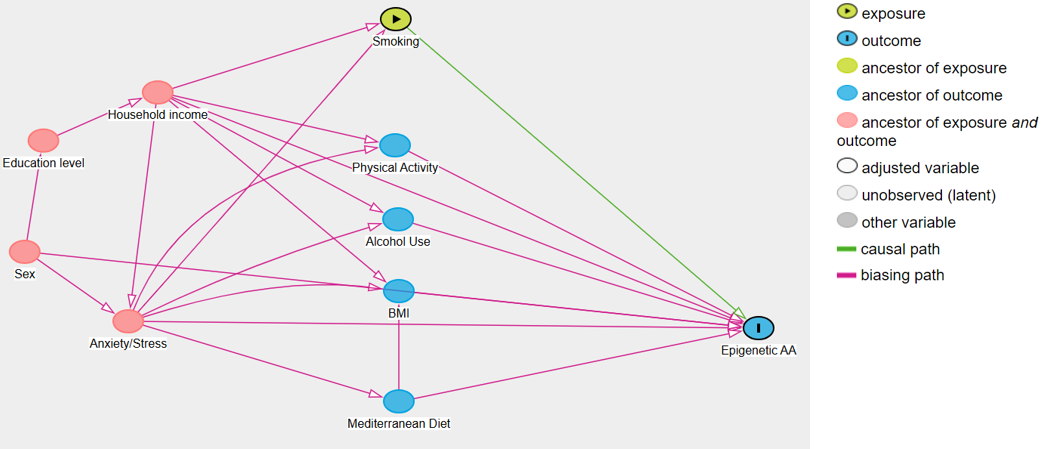

*Figure S2* Smoking is specified as the exposure (green) and EAA as the outcome (blue). Socioeconomic and lifestyle factors, including household income, education level, physical activity, alcohol consumption, body mass index (BMI), and Mediterranean diet, are treated as confounders in the analytical models, as they may influence both smoking behaviour and EAA, although some may also lie on potential mediating pathways. Anxiety/stress is included as a common cause of both smoking and EAA (pink), representing a potential source of confounding. Directed edges represent assumed causal relationships, while highlighted paths indicate potential biasing (non-causal) pathways. Variables adjusted for in the statistical models are indicated by white nodes.

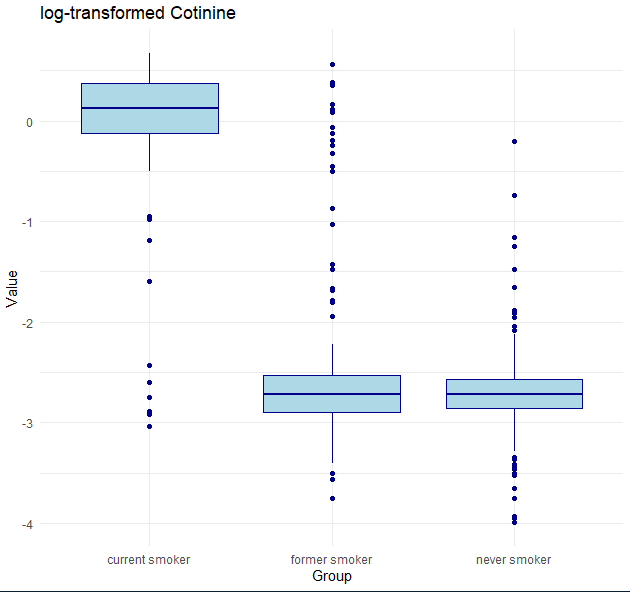

*Figure S3* Boxplot showing the distribution of log-transformed cotinine values by smoking status. Study participants were categorized by smoking status: 98 current smokers, 227 former smokers, and 603 never smokers. The self-reported smoking status was validated using log-transformed cotinine levels. Current smokers (n = 98) had the highest log-transformed cotinine values, followed by former smokers (n = 227), with never smokers (n = 603) exhibiting the lowest values. This validation confirms the accuracy of self-reported smoking status, with clear differences in cotinine levels across the smoking groups. Among the 928 participants, 105 self-reported never smokers had undetectable cotinine levels

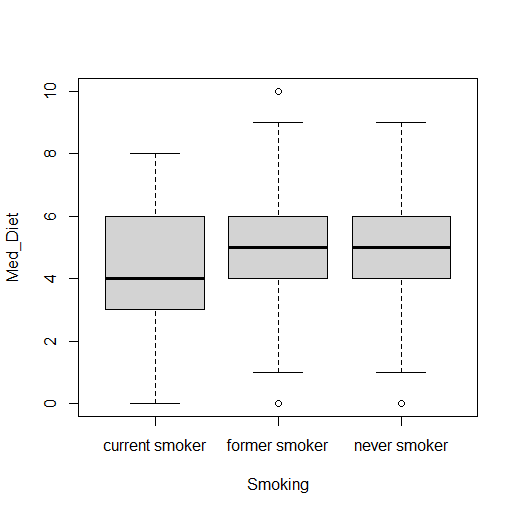

**

*Figure S4* ANOVA on Mediterranean Diet - Never smokers have significantly higher Mediterranean diet scores than current smokers (p = 0.002).

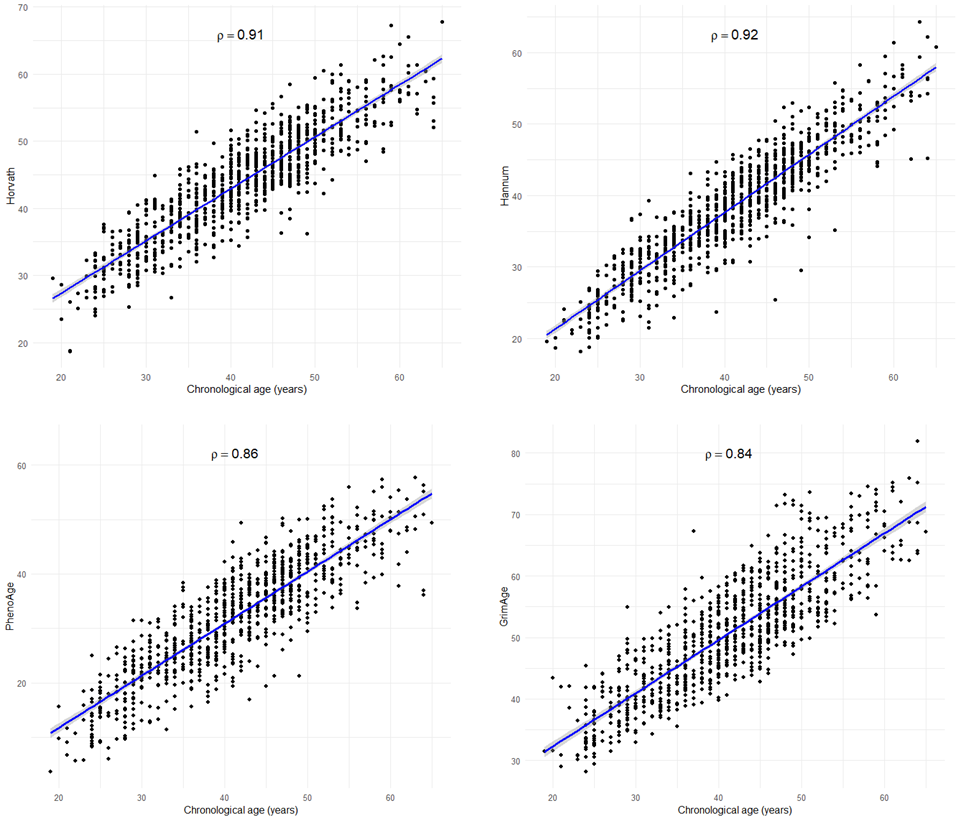

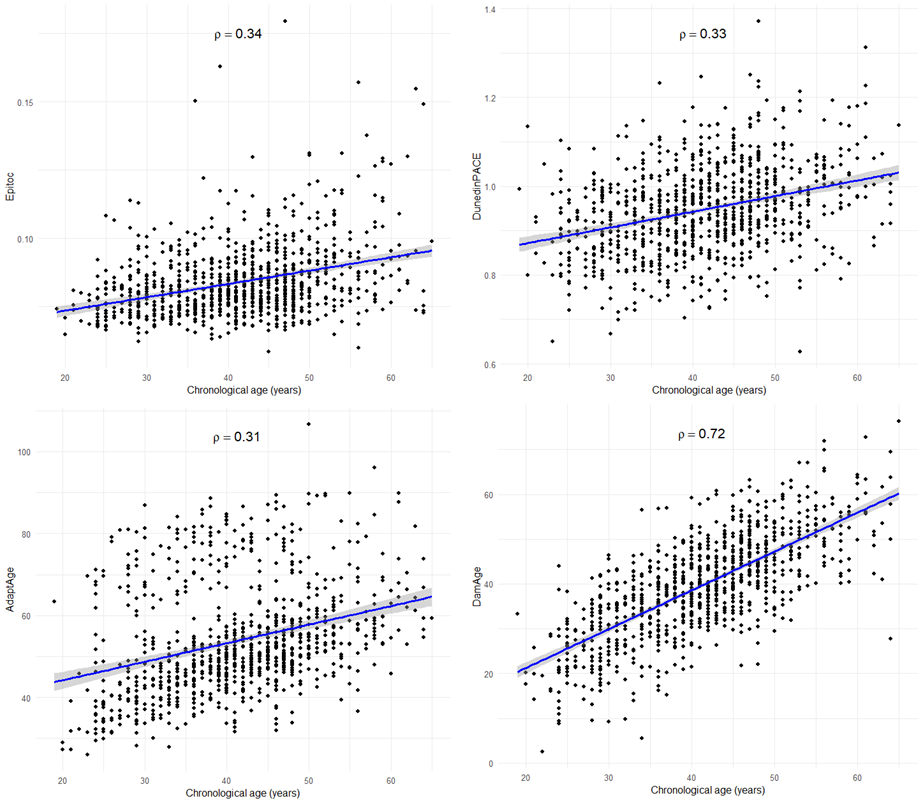

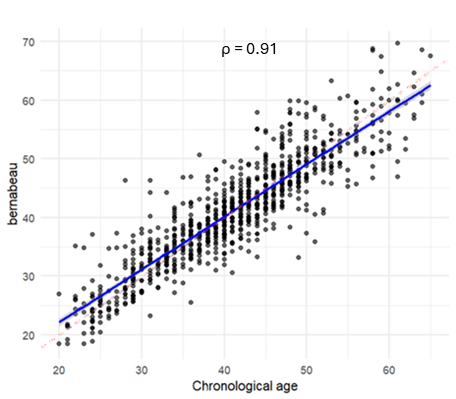

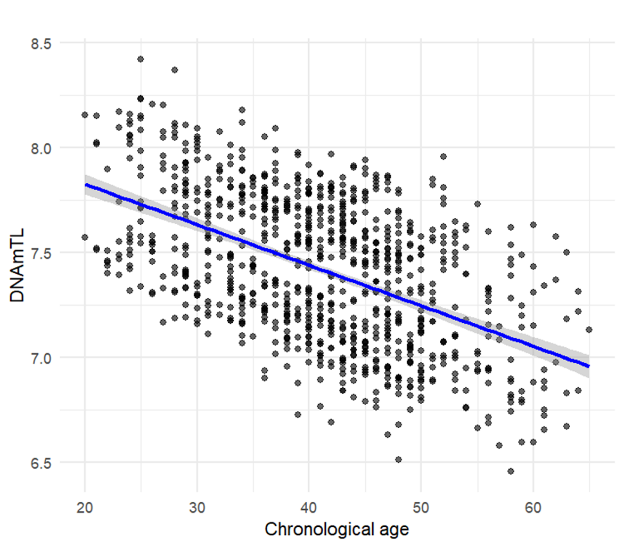

ρ = -0.52

ρ = 0.89

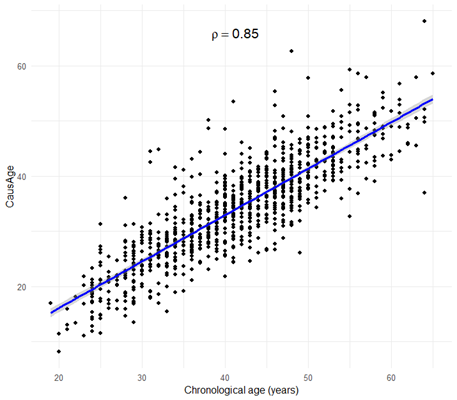

Figure S5 Correlations of raw epigenetic estimates with chronological age (years)
